# Reevaluation of Intravenous Steroid Therapy for Histologically Confirmed Myocarditis with Fulminant Presentation: Insights from the Japanese Registry of Fulminant Myocarditis

**DOI:** 10.1101/2023.09.13.23295517

**Authors:** Hideki Kawai, Hideo Izawa, Masanobu Yanase, Akira Yamada, Hiroshi Takahashi, Yukio Ozaki, Kayoko Takada, Koshiro Kanaoka, Kenji Onoue, Yoshihiko Saito, the Japanese Registry of Fulminant Myocarditis Investigators

## Abstract

**Background:** The efficacy of intravenous steroids (IS) in patients with fulminant myocarditis presentation (FMP) remains controversial. We aimed to compare the clinical outcomes between patients with FMP who received IS (IS(+)) and not received IS (IS(-)).

**Methods:** From the Japanese Registry of Fulminant Myocarditis, we extracted the data of patients requiring catecholamines or mechanical support, with histologically confirmed FMP. The primary outcome was a composite of mortality and heart transplantation within 90 days. We assessed the impact of IS on outcomes using the Kaplan-Meier method, log-rank test, and Cox regression analysis. Patients were categorized according to the number of prognostic factors (age ≥55 years, non-sinus rhythm, left ventricular ejection fraction [LVEF] <40% at admission, ventricular tachycardia/fibrillation on the first day, and the presence of giant cell myocarditis); the relationship between the 90-day prognosis and IS use within these categories.

**Results:** Of 344 patients (median age: 54 years; 40% female), 98 died within 90 days and 16 died after 90 days. IS was administered in 195 patients. The proportion of patients with lymphocytic myocarditis and LVEF were lower in the IS(+) group than in the IS(-) group. Intra-aortic balloon pumping, extracorporeal membrane oxygenation, and intravenous immunoglobulin administration were more common in the IS(+) group than in the IS(-) group. Analysis of the entire cohort indicated worse 90-day outcomes in the IS(+) group than in the IS(-) group (36.3% vs. 19.2%, P=0.0021); however, there was no substantial difference after propensity score matching (PSM; 26.2% vs. 24.2%; P=0.95). On unadjusted Cox regression, IS use was associated with worse 90-day outcomes (hazard ratio, 1.95 [95% confidence interval, 1.26-3.04]; P=0.0026). However, after PSM, this association was no longer significant (1.02 [0.56-1.87], P=0.95). Similar results were observed among patients with lymphocytic myocarditis. The prognosis was notably worse with IS administration than without IS administration among low-risk patients (P=0.001).

**Conclusions:** IS may not provide prognostic advantages for patients with FMP. The adverse effects of IS might be more pronounced in low-risk patients.

**Registration:** URL: https://www.umin.ac.jp/ctr; Unique identifier: UMIN000039763.

**Clinical Perspective:** *What Is New?:*

- Intravenous steroids were commonly administered in more severe cases, particularly in patients diagnosed with eosinophilic myocarditis or giant cell myocarditis.
- While the prognosis was poorer in patients who received intravenous steroids than in those who did not, the outcomes were similar when comparing cohorts matched on patient background factors.
- Notably, prognosis was worse in low-risk patients who were administered intravenous steroids.

*What Are the Clinical Implications?:*

- Administering intravenous steroids might not yield any prognostic advantage for patients with fulminant myocarditis. Further, the potential negative effects of intravenous steroids appear to be more pronounced in low-risk patients.
- Thus, clinicians should be cautious about administering intravenous steroids, especially in those identified as low-risk.

## Introduction

Fulminant myocarditis presentation (FMP), an uncommon syndrome characterized by sudden and severe cardiac inflammation, requiring inotropes or mechanical circulatory support, is associated with an extremely high risk of death due to cardiogenic shock, fatal ventricular tachyarrhythmias, or bradyarrhythmia.^1–4^ The early recognition of FMP, introduction of inotropes and/or mechanical circulatory support, and maintenance of end-organ function can contribute to favorable outcomes. However, the efficacy of immunosuppressive therapy for FMP, in addition to intensive care management, remains controversial.

Although a few prospective trials on immunosuppressive therapy for lymphocytic myocarditis (LM) have been performed, they essentially targeted the chronic disease course and not the acute inflammatory phase. In cases of fulminant disease and cardiogenic shock, corticosteroids are often used, despite the lack of clear evidence of benefits and an undefined risk of adverse effects. Currently, no evidence is available to support the efficacy of immunosuppressive therapy for acute LM.^3,4^ Eosinophilic myocarditis (EM) has been reported to respond well to steroid therapy in specific cases, although large-scale evidence is lacking.^5–7^ Symptoms may be resolved with concomitant immunosuppressants, even in patients who require inotropes or mechanical circulation assistance.^8,9^ Additionally, the prognosis of patients with giant-cell myocarditis (GCM) is improved with immunosuppressive therapy; specifically, the survival rate is not improved with steroids alone, but is improved with a combination of steroids and other immunosuppressants.^10,11^

Given the above-mentioned limited data, this multicenter cohort study aimed to determine the prognostic efficacy of intravenous steroids (IS) in patients with FMP using data from the Japanese Registry of Fulminant Myocarditis.^12^

## Methods

### Study Design and Population

The Japanese Registry of Fulminant Myocarditis is a multicenter, retrospective cohort study conducted in collaboration with 235 cardiovascular training hospitals in Japan, as described in a previous report.^12^

Using the Japanese Registry of All Cardiac and Vascular Diseases–Diagnosis Procedure Combination discharge database, which includes a claims database covering >60% of all cardiovascular training hospitals in Japan,^13,14^ we extracted with data of patients diagnosed with myocarditis between April 2012 and March 2017 using International Classification of Diseases–10 codes, I40, I41, and I423. We categorized patients with myocarditis into FMP and non-FMP groups based on catecholamine or mechanical support use during hospitalization.^15,16^ Individual patient data (retrospective anonymous data) were collected from each facility that joined the registry, with approval obtained from the local institutional review board. Of 501 facilities screened across Japan, 235 joined the registry and registered data. The cardiologists at each participating center manually reviewed the charts to extract individual data. Treatment details during hospitalization were extracted from the claims database. The data were uploaded into the Research Electronic Data Capture web app (Vanderbilt University, TN, USA). Data quality and logical errors were centrally checked by Drs. Kanaoka and Sumita; when needed, local investigators were contacted for clarification or details of the data.

Histologically proven myocarditis was defined according to the clinical diagnostic criteria of the European Society of Cardiology or the Japanese Circulation Society and the histologic definitions of World Health Organization/International Society and Federation of Cardiology criteria.^3,17,18^ Patients were excluded if they were diagnosed with other diseases, symptoms appeared more than 30 days before admission, or catecholamine or mechanical circulatory support were started after the 14th day of hospitalization. Patients with a recorded day of IS introduction were defined as the IS(+) group, and those in whom IS were never administered were defined as the IS(-) group.

Follow-up data were obtained from medical records or telephone interviews. The primary outcome was a composite of all-cause mortality or heart transplantation (HTx) within 90 days of follow up; long-term all-cause mortality or HTx was also analyzed. Based on a previous study using the same registry, older age (≥55 years), non-sinus rhythm, low left ventricular ejection fraction (LVEF; <40%) on admission, and ventricular tachycardia or fibrillation on the day of admission were considered as prognostic factors.^12^

The study protocol was approved by the Ethics Committee of Nara Medical University (registration number: 2256) in July 2019 and by the Ethics Committee of the Japanese Circulation Society (registration number: 10) in November 2019. This study was conducted in accordance with the principles of the Declaration of Helsinki. The data, analytical methods, and study materials are not available to other researchers to reproduce the results or replicate the procedure.

### Statistical Analysis

The Shapiro-Wilk test was used to assess the normality of continuous data. Variables with a normal distribution are expressed as the mean ± standard deviation, and asymmetrically distributed data are presented as the median and interquartile range. Categorical variables are presented as frequencies (percentages). Differences between groups were evaluated using the Mann–Whitney U test or Student’s t-test for continuous variables and the chi-squared test for categorical variables.

We compared patient characteristics between IS(+) and IS(-) groups in accordance with previous studies.^12^ To control for potential confounders, we conducted propensity score matching (PSM), in both the entire and LM populations. PSM was performed using a greedy matching algorithm with a multivariate logistic regression model that included all baseline variables. We assessed the impact of IS on 90-day and long-term outcomes using the Kaplan-Meier method, log-rank test, and Cox hazard analysis, in the crude and PSM populations. Additionally, the impact of IS on the 90-day prognosis was examined according to the number of prognostic factors, as well as according to the timing of IS initiation. The formal interaction test was performed between the subgroup according to the number of prognostic factors and the effects of IS.

All statistical tests were two-sided and P-values <0.05 were considered statistically significant. SPSS 22 (SPSS Inc, Chicago, IL, USA) was used for all statistical analyses.

## Results

### Study Population

Of the 344 patients (median age: 54 years; 40% female) with histologically confirmed FMP during the study period, 98 died within 90 days and 16 died after 90 days. IS was administered in 195 patients (IS(+) group), and not in the remaining 149 (IS(-) group). The LVEF level and proportion of patients with LM were significantly lower in the IS(+) group than in the IS(-) group. In contrast, the use of intra-aortic balloon pumping, extracorporeal membrane oxygenation, and intravenous immunoglobulin was more frequent in the IS(+) group than in IS(-) group. After PSM using all baseline characteristics shown in Table 1, the differences between IS(+) and IS(-) groups were no longer significant. Similarly, among patients with LM by endomyocardial biopsy (n=273), there were significant differences between IS(+) and IS(-) groups in LVEF and the use of intra-aortic balloon pumping, extracorporeal membrane oxygenation, ventricular assist device, and intravenous immunoglobulin in the crude population, but not after PSM (Table 2).

**Table 1.**
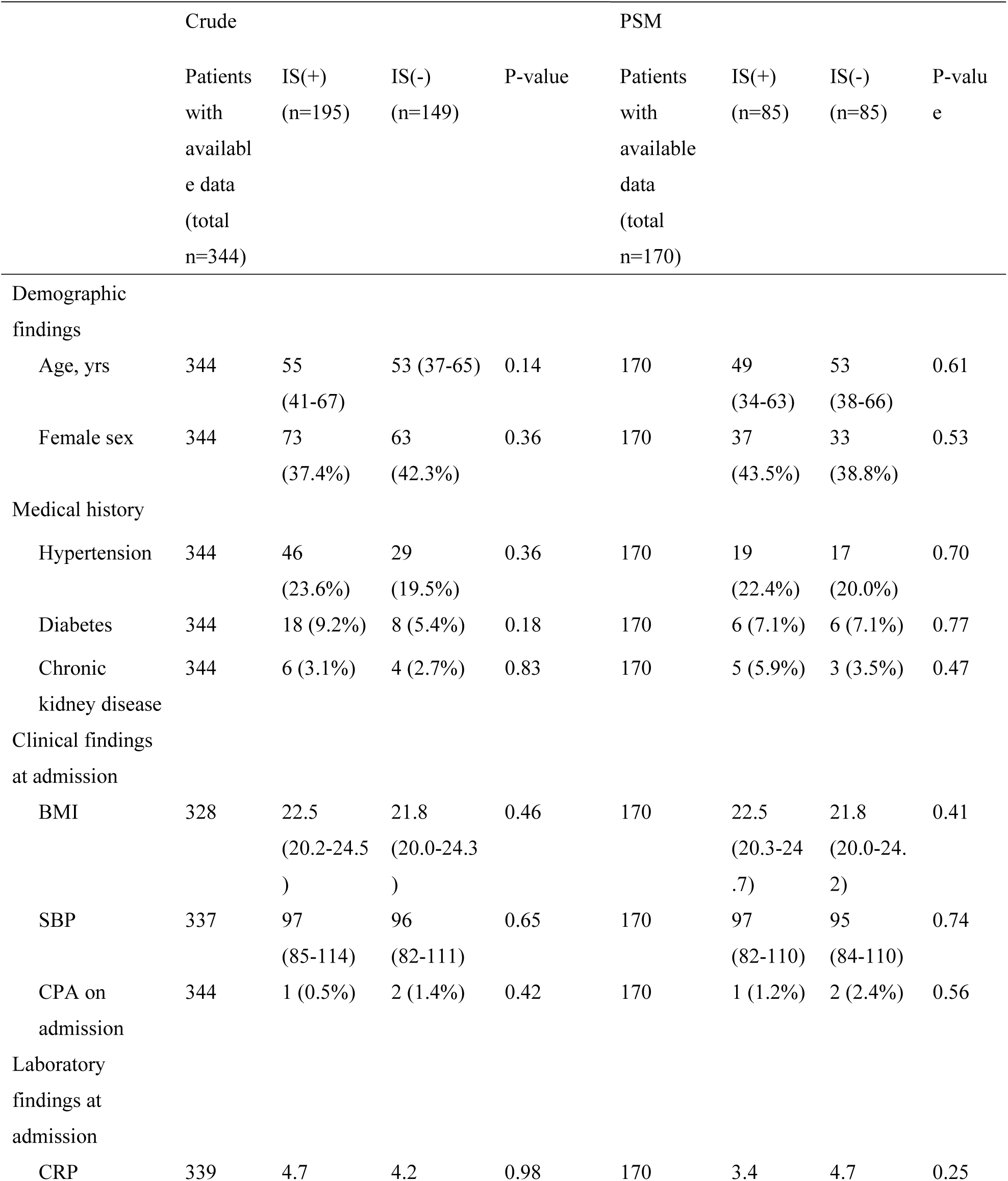
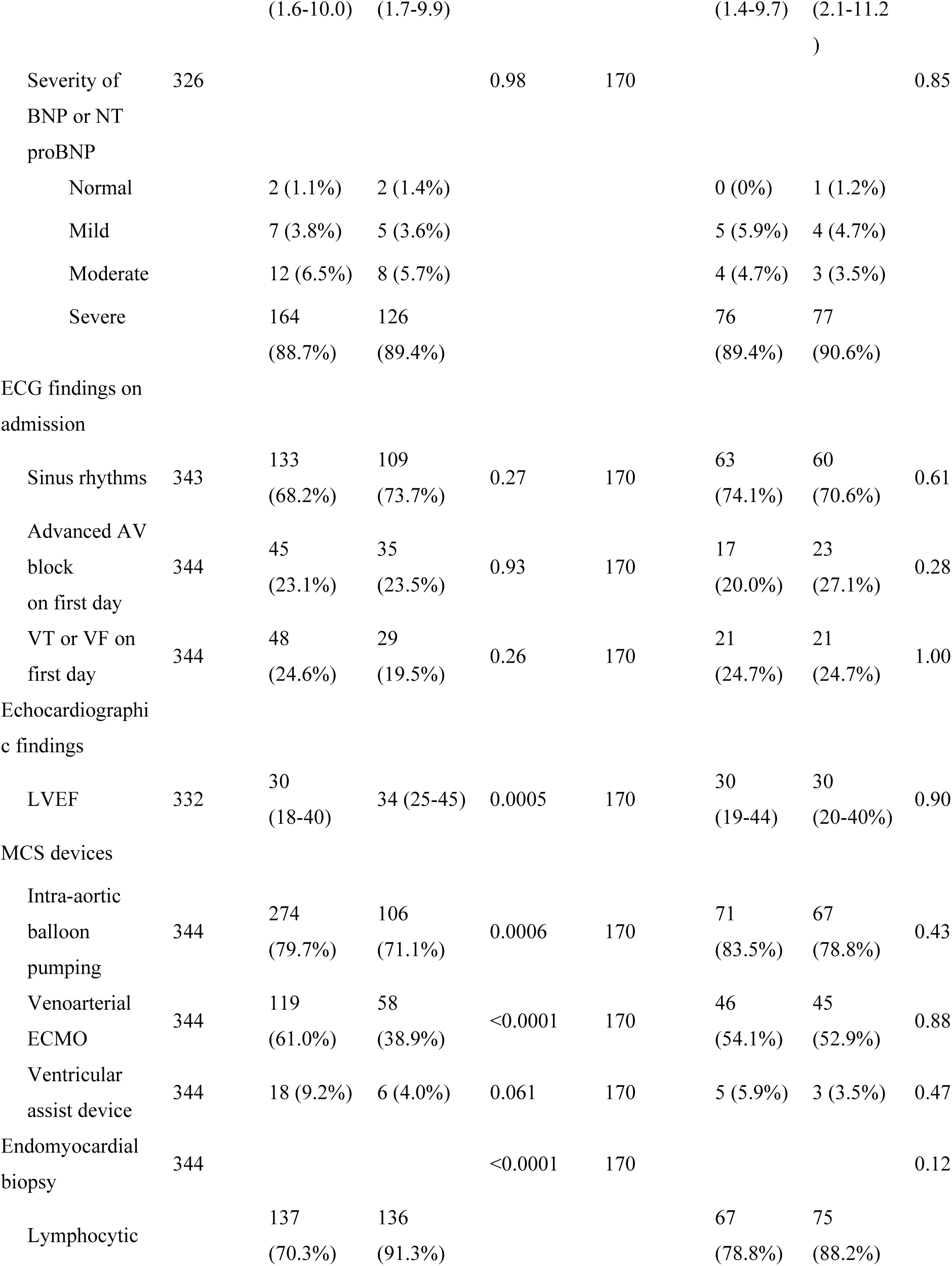
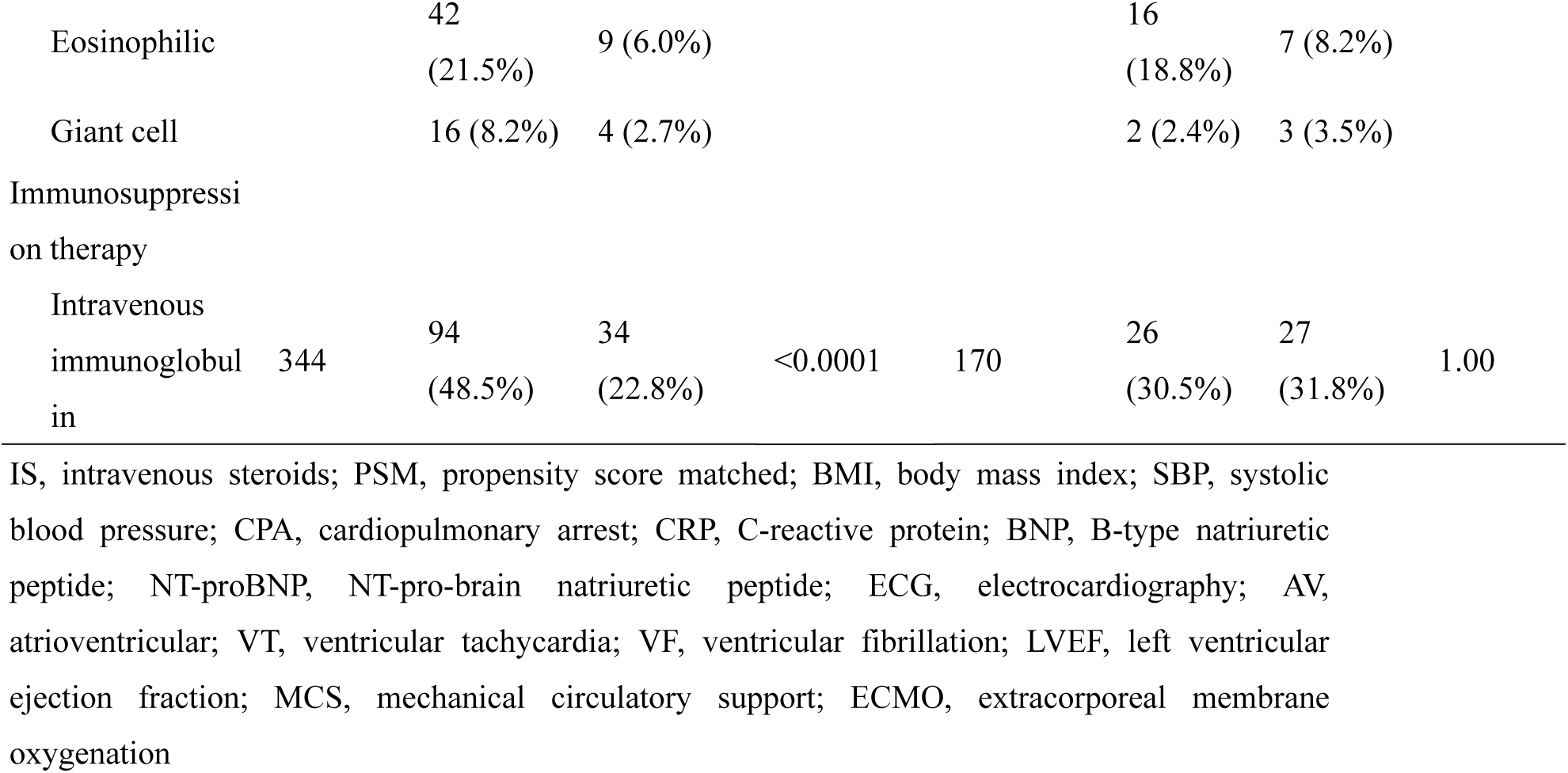
Baseline characteristics of patients with fulminant myocarditis who did and did not receive IS.

**Table 2.**
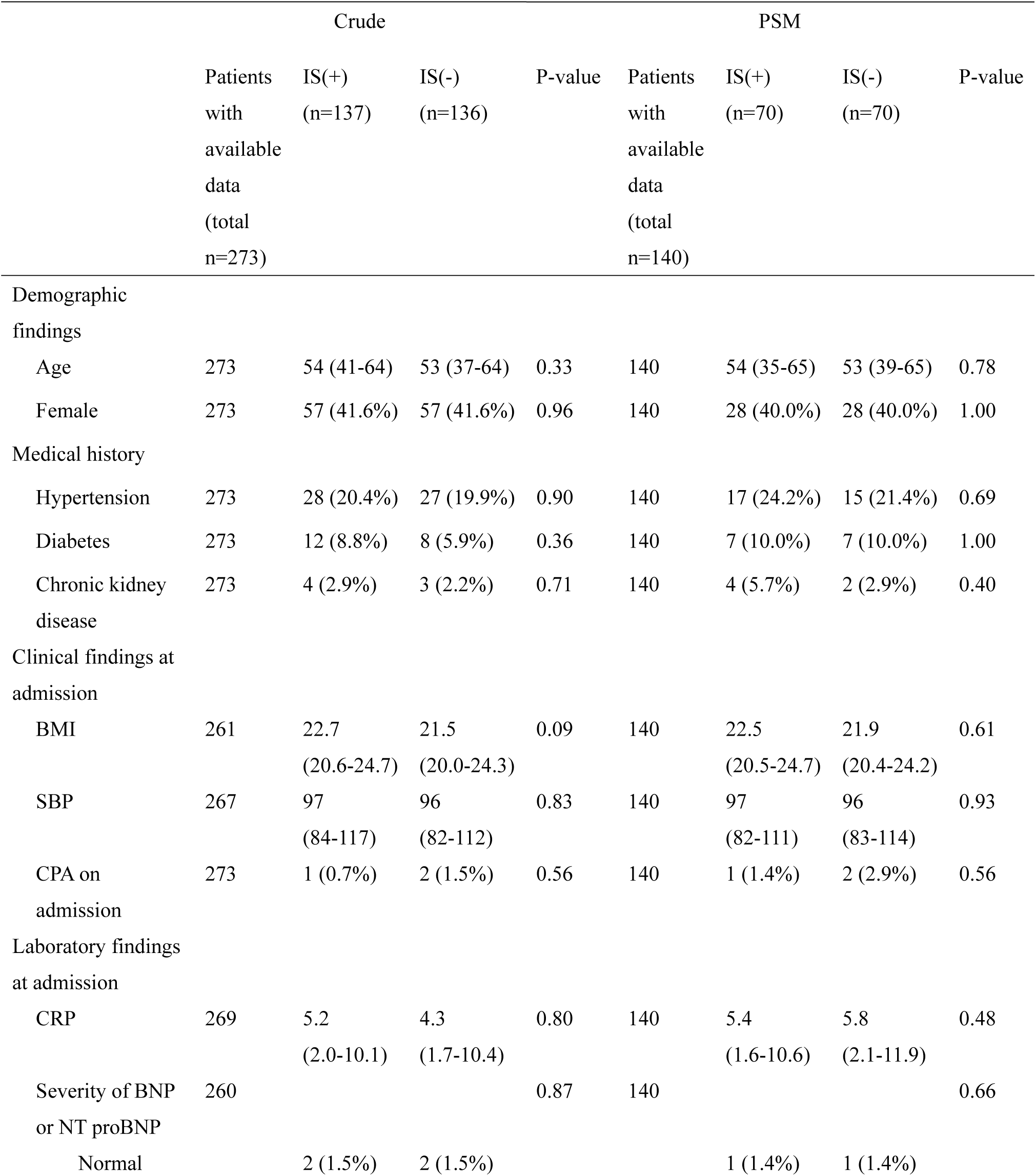
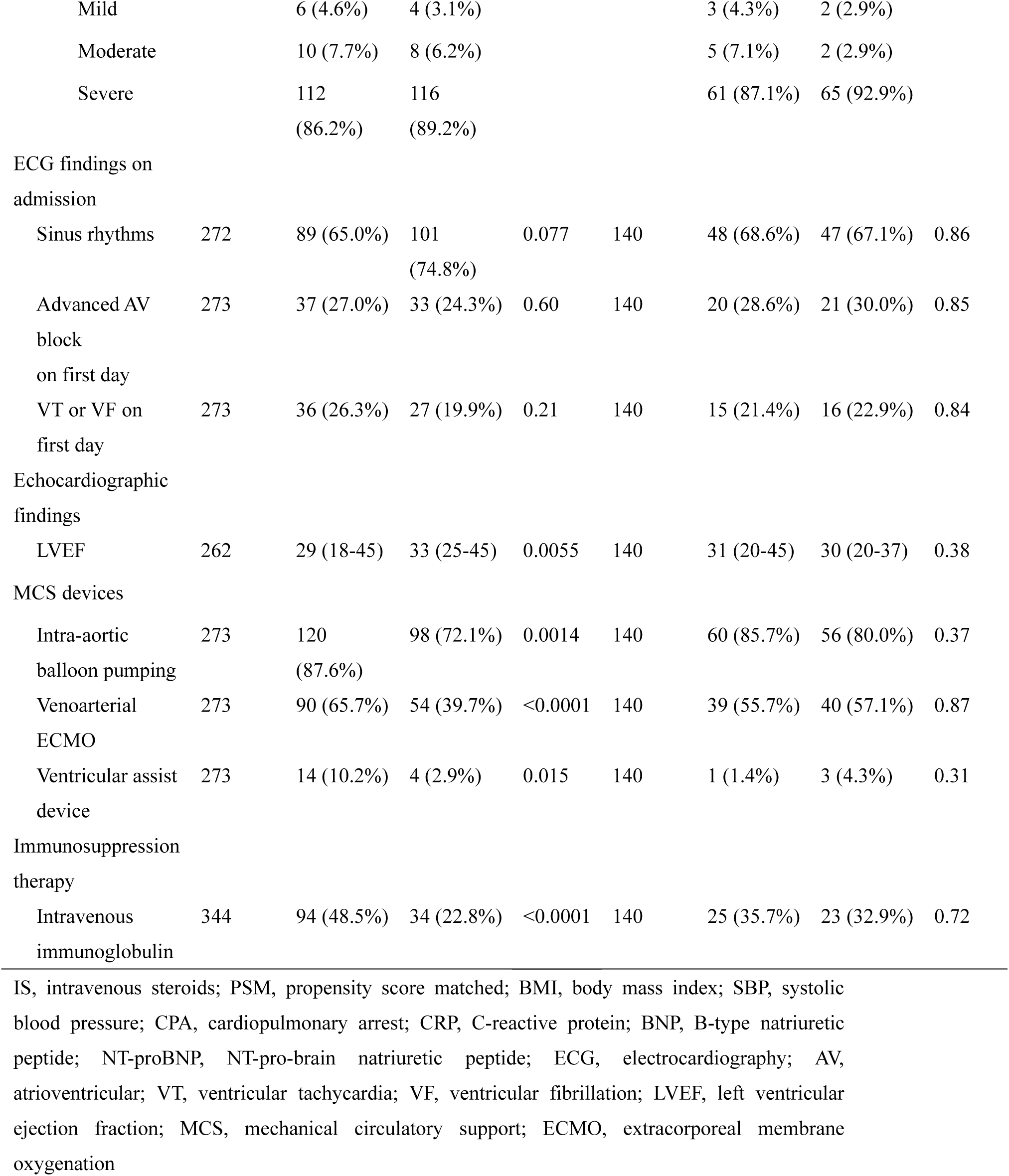
Baseline characteristics of patients with lymphocytic fulminant myocarditis who did and did not receive IS.

### Clinical Outcomes

The outcomes of all fulminant myocarditis subtypes are shown in Figure 1. Of the 344 patients, 70 of 195 patients in the IS(+) group and 28 of 149 patients in the IS(-) died or underwent HTx within 90 days. Additionally, 80 of 195 patients in the IS(+) group and 34 of 149 patients in the IS(-) group died or underwent HTx after 90 days. Although the crude analysis showed worse 90-day (36.3% vs. 19.2%, P=0.0021) and long-term outcomes (42.3% vs. 22.9% at 3 years, P=0.0016) in the IS(+) group than in the IS(-) group, there was no significant difference in the PSM analysis (26.2% vs. 24.2%, P=0.95 for 90-day outcomes; and 31.1% vs. 28.8% at 3 years, P=0.92 for long-term outcomes).

**Figure 1.**
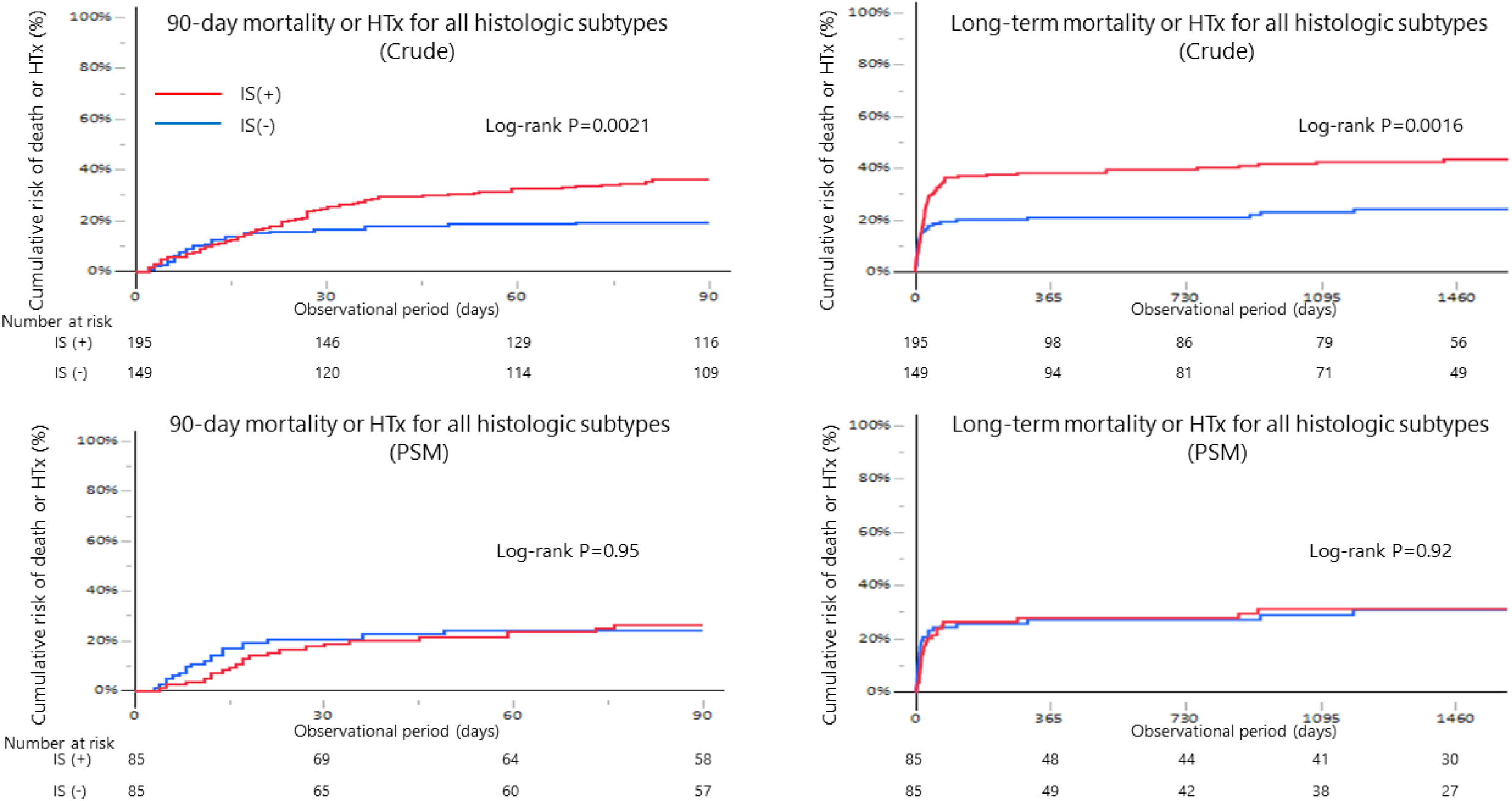
Association of IS use with prognosis in patients with fulminant myocarditis (all histologic subtypes) (a) 90-day mortality or HTx (Crude) (b) Long-term mortality or HTx (Crude) (c) 90-day mortality or HTx (PSM) (d) Long-term mortality or HTx (PSM) IS, intravenous steroid; HTx, heart transplantation; PSM, propensity score matched

The prognosis of patients with fulminant LM is shown in Figure 2. Of the 273 patients, 47 of 137 patients in the IS(+) group and 26 of 136 patients in the IS(-) group died or underwent HTx within 90 days. Additionally, 54 of 137 patients in the IS(+) group and 31 of 136 patients in the IS(-) group died or underwent HTx after 90 days. Although the crude analysis showed worse 90-day (34.7% vs. 19.6%, P=0.0140) and long-term outcomes (40.3% vs. 22.9% at 3 years, P=0.0100) in the IS(+) group than in the IS(-) group, there were no significant differences in the PSM analysis (30.4% vs. 26.3%, P=0.70 for 90-day outcomes; and 32.5% vs. 30.3% at 3 years, P=0.94 for long-term outcomes).

**Figure 2.**
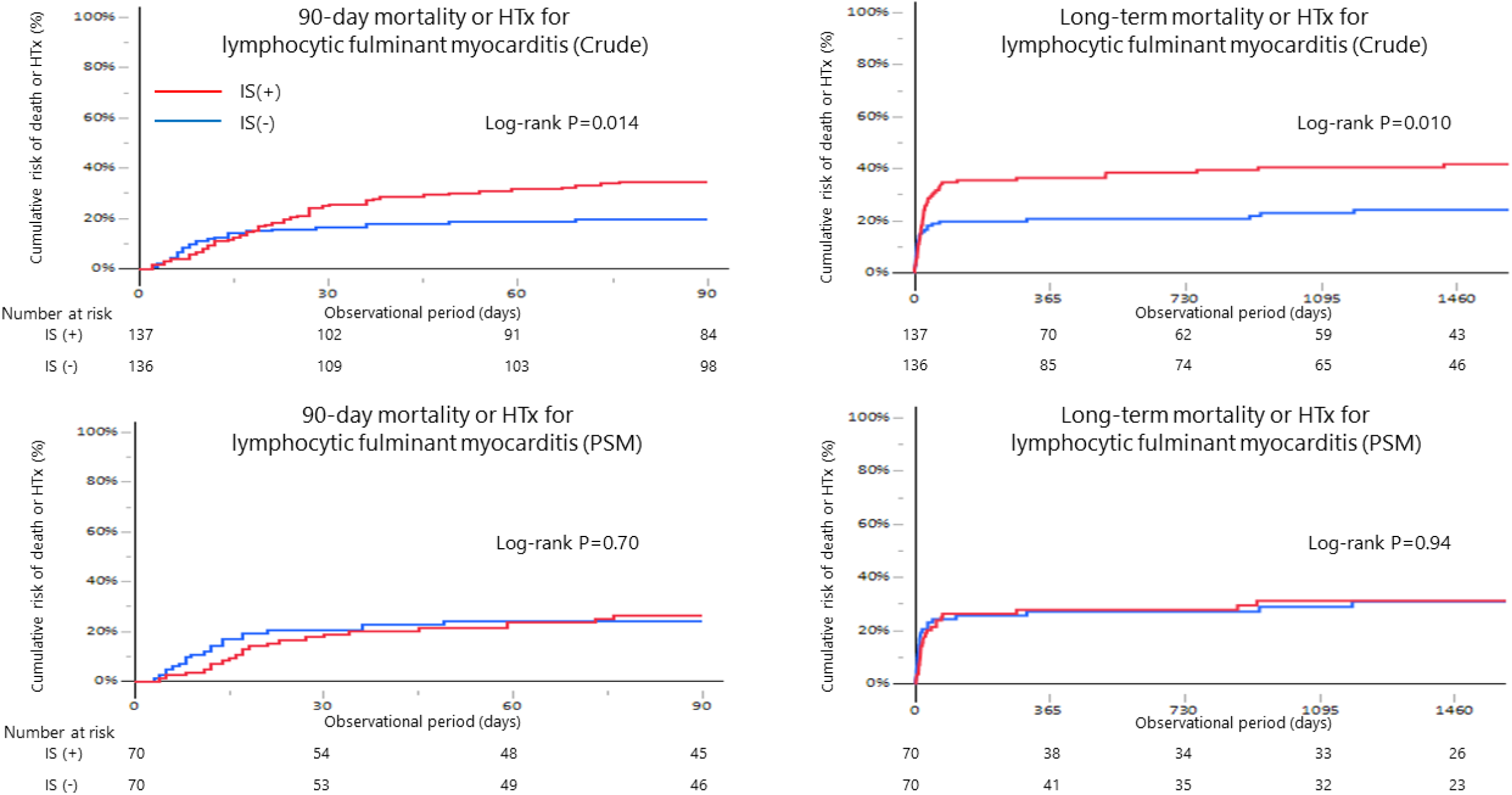
Association of IS use with prognosis in patients with lymphocytic fulminant myocarditis (a) 90-day mortality or HTx (Crude) (b) Long-term mortality or HTx (Crude) (c) 90-day mortality or HTx (PSM) (d) Long-term mortality or HTx (PSM) IS, intravenous steroid; HTx, heart transplantation; PSM, propensity score matched

The prognoses of patients with fulminant EM and GCM are shown in Supplemental Figure S1. In the analysis of only patients with EM, the differences did not reach significance because of the small number of cases; however, a tendency of a poorer prognosis in the IS(+) group than in the IS(-) group was observed (31.3% vs. 11.1%, P=0.26 for 90-day outcomes; and 37.7% vs. 22.2% at 3 years, P=0.42 for long-term outcomes). Similarly, in the analysis of patients with GCM, there were also no significant differences in outcomes between the IS(+) and IS(-) groups (62.3% vs. 25.0%, P=0.26 for 90-day outcomes, and 70.0% vs. 25.0% at 3 years, P=0.20 for long-term outcomes).

Cox hazard analysis of IS use for 90-day and long-term outcomes was performed for all subtypes and LM. On unadjusted Cox regression, IS use was associated with worse 90-day (hazard ratio, 1.95 [95% confidence interval (CI), 1.26-3.04]; P=0.0026) and long-term outcomes (1.89 [1.26-2.82]; P=0.0020). However, after PSM, the association was no longer significant (1.02 [0.56-1.87], P=0.95 for 90-day outcomes; and 0.97 [0.56-1.70], P=0.92 for long-term outcomes). Similarly, among patients with LM, IS use was associated with worse 90-day (hazard ratio, 1.80 [95% CI, 1.12-2.91], P=0.016) and long-term outcomes (1.77 [1.14-2.76], P=0.011) in the crude analysis. However, after PSM, this association was no longer significant (1.13 [0.60-2.13], P=0.70 for 90-day outcomes; and 1.02 [0.56-1.86], P=0.94 for long-term outcomes) (Table 3).

**Table 3.** Cox hazard analysis of the impact of intravenous steroids on 90-day and long-term mortality or HTx.

|  | 90-day mortality or HTx |  | Long-term mortality or HTx |  |
| --- | --- | --- | --- | --- |
|  | HR (95% CI) | P-value | HR (95% CI) | P-value |
| All Subtypes |  |  |  |  |
| Crude | 1.95 (1.26-3.04) | 0.0026 | 1.89 (1.26-2.82) | 0.0020 |
| PS adjusted | 1.09 (0.63-1.90) | 0.76 | 1.01 (0.61-1.68) | 0.96 |
| PS matched | 1.02 (0.56-1.87) | 0.95 | 0.97 (0.56-1.70) | 0.92 |
| Lymphocytic |  |  |  |  |
| Crude | 1.80 (1.12-2.91) | 0.016 | 1.77 (1.14-2.76) | 0.011 |
| PS adjusted | 1.02 (0.55-1.89) | 0.95 | 0.95 (0.54-1.66) | 0.85 |
| PS matched | 1.13 (0.60-2.13) | 0.70 | 1.02 (0.56-1.86) | 0.94 |

HTx, heart transplantation; HR, hazards ratio; CI, confidence interval; PS, propensity score

### Analysis According to the Number of Prognostic Risk Factors

In a previous study using the same registry, factors such as older age, non-sinus rhythm, low LVEF (<40%) upon admission, and presence of ventricular tachycardia or fibrillation on the day of admission were identified as predictors of reduced 90-day survival.^12^ In the current study, we compared outcomes between IS(+) and IS(-) groups, stratified by the number of previously identified risk factors. Although minimal differences in 90-day mortality rates were observed between IS(+) and IS(-) groups among patients with 2, 3, and 4 or more risk factors, the 90-day mortality rate was notably higher in the IS(+) group than in the IS(-) group among patients with 0 or 1 factors (i.e. low-risk patients) (25% vs 1%) in the crude analysis (P=0.001 for the interaction) (Figure 3a). This difference was also significant in the LM population (Figure 3b), but not in the EM and GCM populations (Figure 3c).

**Figure 3.**
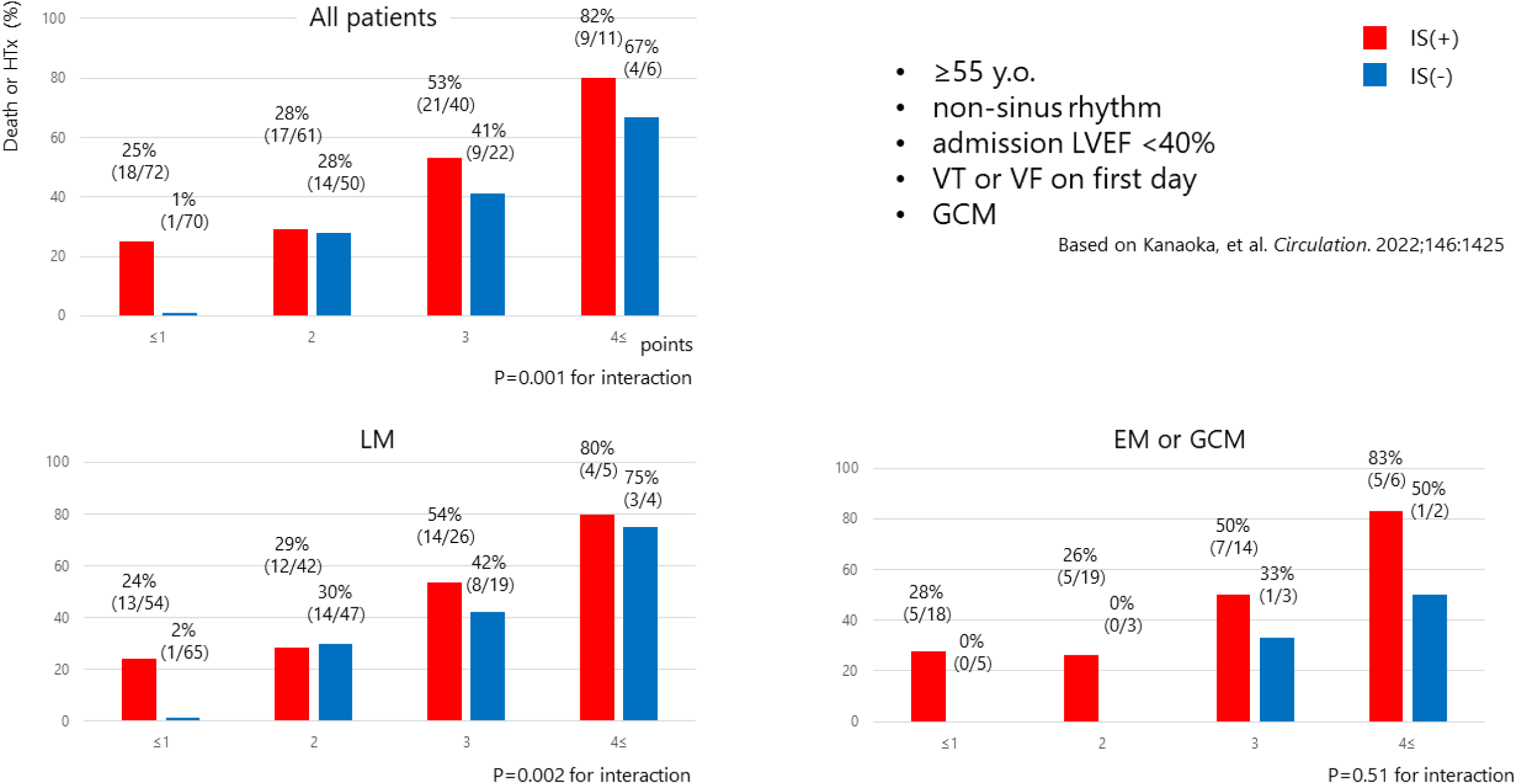
Steroid administration versus non-administration across various risk stratifications LVEF, left ventricular ejection fraction; VT, ventricular tachycardia; VF, ventricular fibrillation; LM, lymphocytic myocarditis; EM, eosinophilic myocarditis; GCM, giant cell myocarditis; HTx, heart transplantation

### Analysis According to IS Initiation Timing

Therapeutic strategy adjustments based on the number of days from admission are shown in Figure 4a. On the first day of admission, 54 patients started IS treatment; among the remaining 290 patients, IS was subsequently introduced in 43 patients on day 2, 23 on day 3, 20 on day 4, 16 on days 5-7, and 39 on day 8. Modifications to the therapeutic approach based on the number of days following endomyocardial biopsy (EMB) are illustrated in Figure 4B. IS was introduced either before or within two days after EMB in 142 patients; among the remaining 202 patients, IS was subsequently initiated in 10 patients on day 3, 11 on days 4-5, 6 on days 6-7, and 26 on or after day 8 post-EMB.

**Figure 4.**
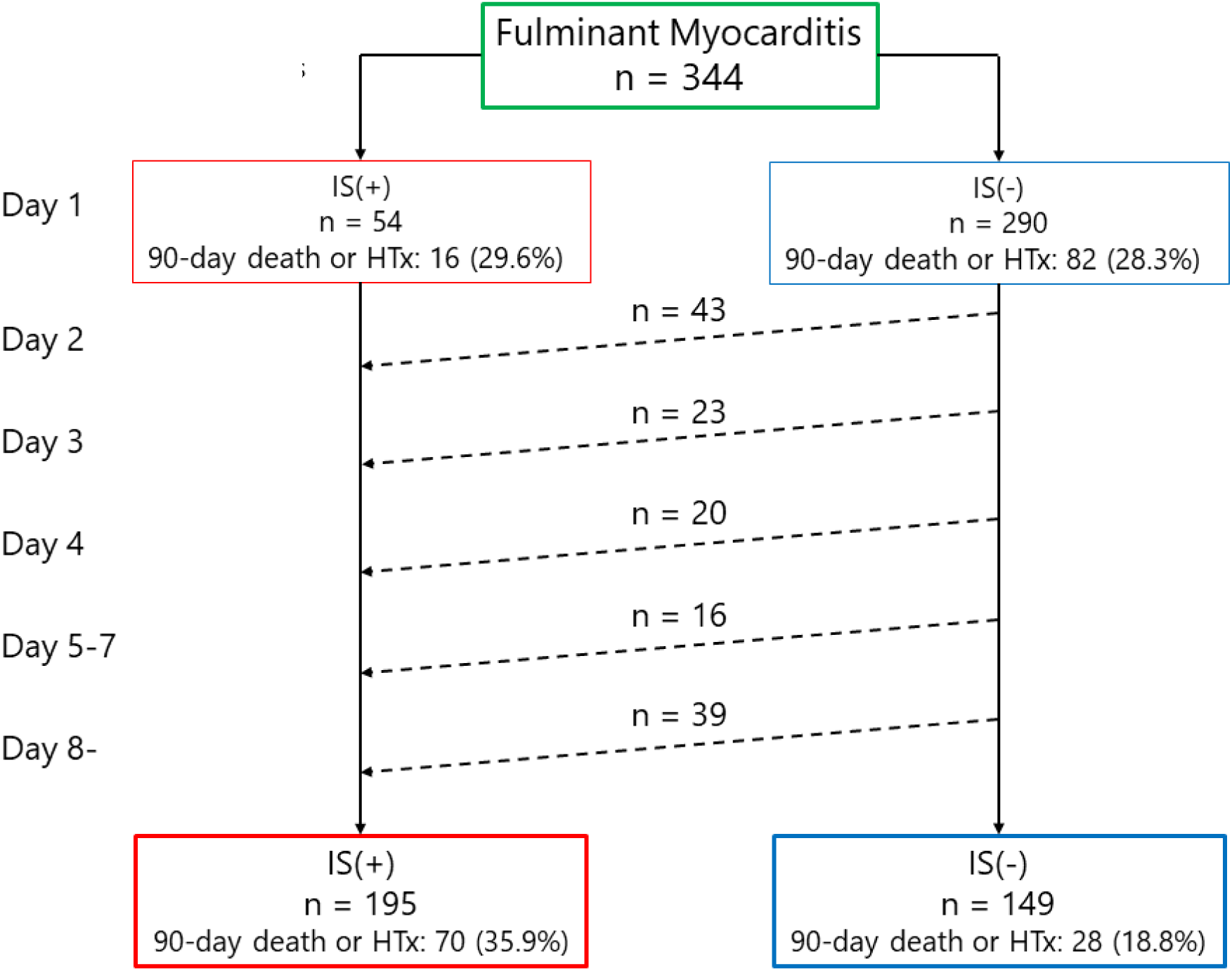
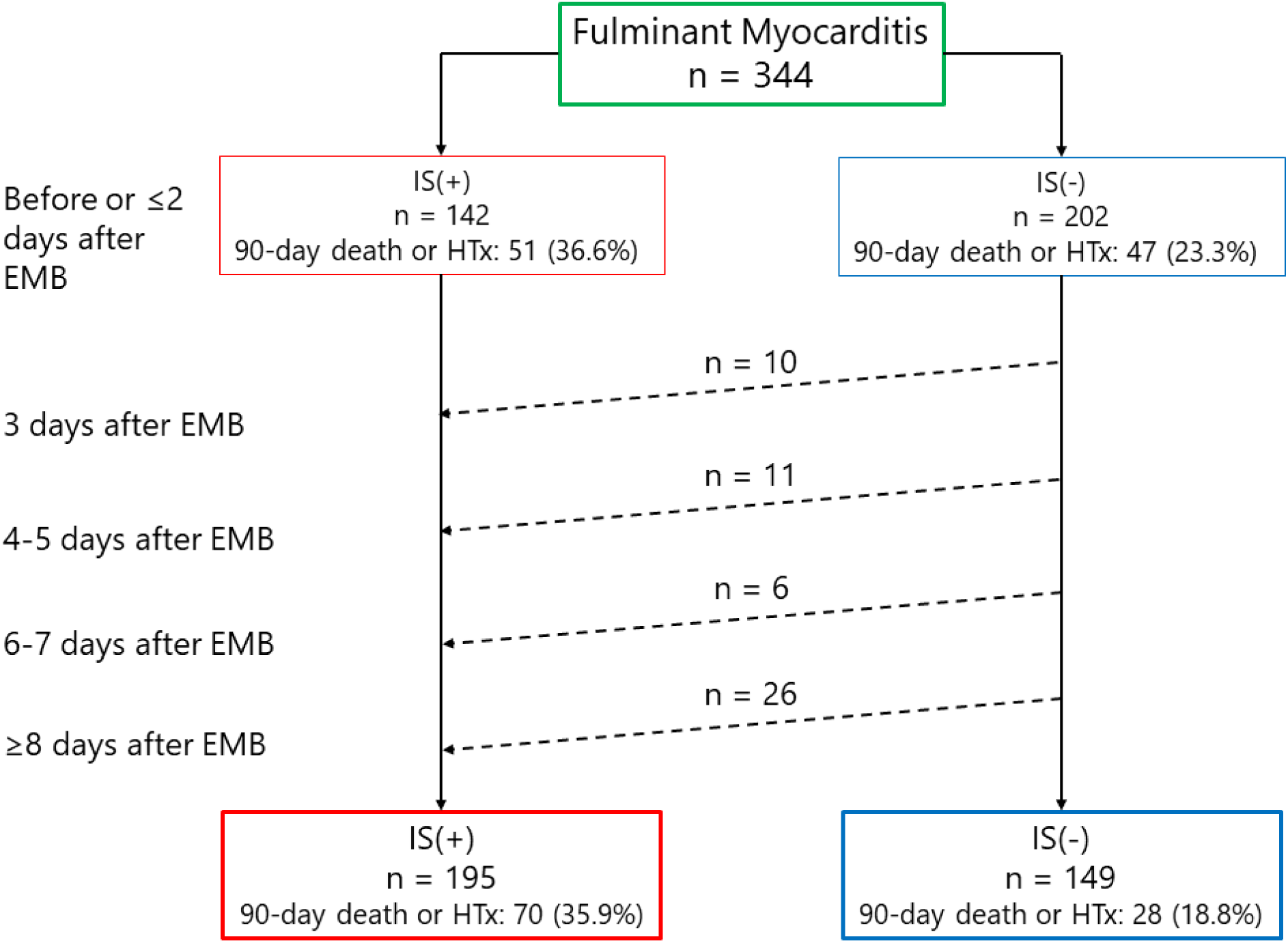
IS initiation patterns **a.** IS initiation based on the number of days after admission. **b.** IS initiation based on the number of days after endomyocardial biopsy. IS, intravenous steroid; HTx, heart transplantation

Figure 5 shows the association between the IS initiation timing and outcomes. The rate of 90-day mortality or HTx was 29.6% (16 of 54) among patients who started IS immediately (on day 1), 38.3% (54 of 141) among those who started IS on day 2 or later, and 18.8% (28 of 149) among those who did not receive IS.

**Figure 5.**
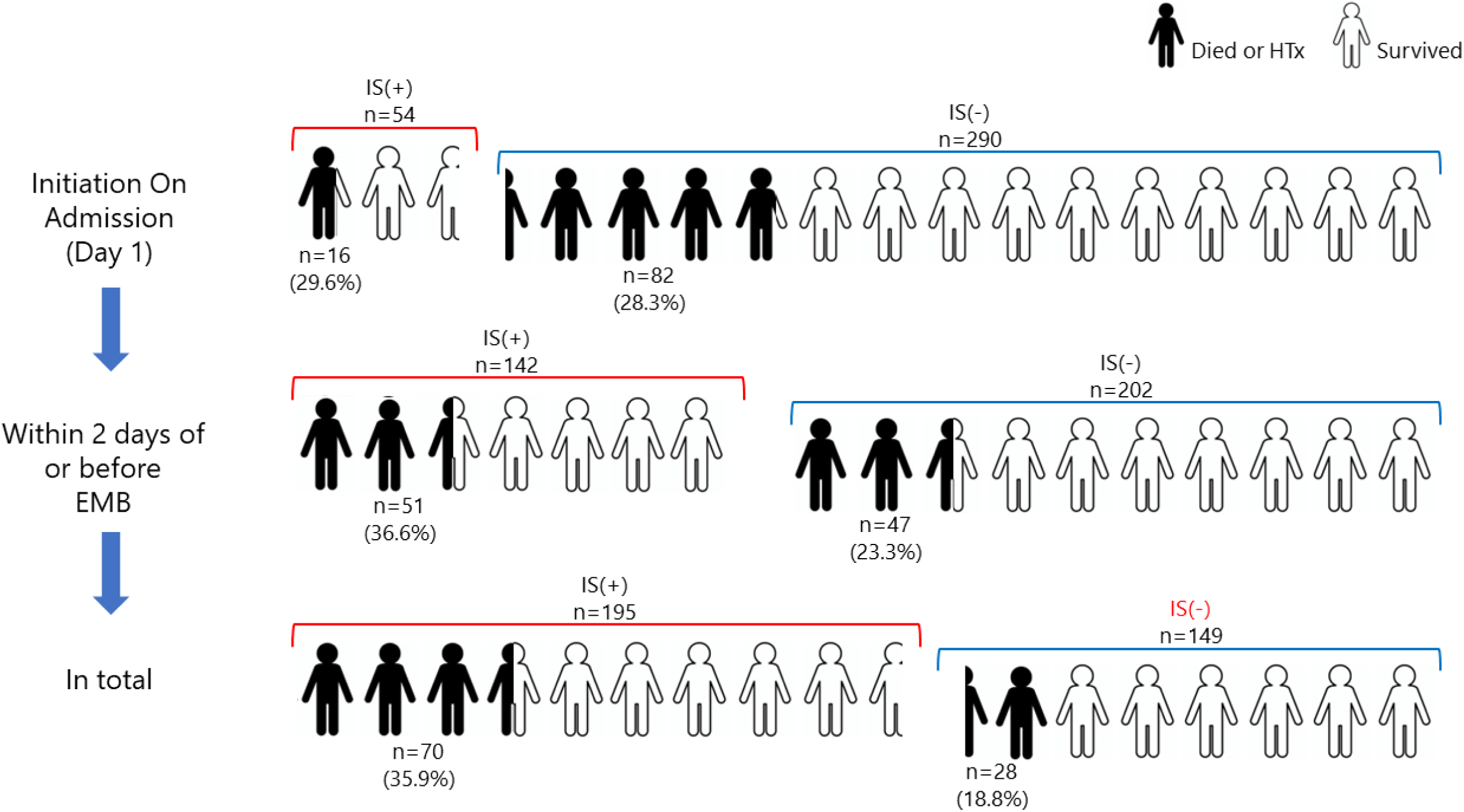
Association between IS initiation timing and outcomes IS, intravenous steroid; EMB: endomyocardial biopsy; HTx, heart transplantation

## Discussion

In our cohort of patients with FMP, IS was initiated in approximately half of the patients with LM and in over 80% of the patients with EM or GCM. Notably, patients who were administered IS had lower LVEF upon admission and increased utilization of mechanical circulatory support, such as an intra-aortic balloon pump or venoarterial extracorporeal membrane oxygenation, and immunoglobulin therapy than their counterparts who did not receive IS. While the overall prognosis appeared worse for patients with FMP who received IS than for those who did not, this disparity was no longer significant after PSM based on patient characteristics. These data suggest a higher prevalence of high-risk patients in the IS(+) group than in the IS(-) group, which could contribute to the poorer outcomes. Furthermore, it is particularly noteworthy that low-risk patients with FMP who received IS exhibited a worse prognosis than their IS(-) counterparts. In contrast, the difference in prognosis between the two groups was marginal among high-risk patients with FMP. These results raise concerns that while IS might not offer overall benefits for managing FMP in the entire population, it may also have detrimental effects, especially in low-risk patients with FMP.

In our investigation of 344 patients diagnosed with FMP by EMB, including 273 with LM, 51 with EM, and 20 with GCM, IS was used in 195 (57%) patients. In a study conducted in Italy involving 187 patients, steroids were administered to 43% (23/55) of patients with FMP and 8% (10/132) of those without FMP.^8^ Another review encompassing 220 cases from the United States, European Union, and Japan in which EMB was performed found that steroids were administered to 32.5% (53/165) of patients in the FMP group and 18.3% (15/55) of patients in the non-FMP group.^15^ Additionally, a report from Italy detailed 443 cases in which EMB was performed in 56 patients (38 with FMP).^19^ Although this report did not specify steroid usage, the relatively low percentage of patients with FMP suggests that steroid administration was likely limited. Notably, although there have been multiple FMP registry reports, our study is the first to compare outcomes between patients with FMP who did and did not receive steroid treatment.

Furthermore, we retrospectively classified patients with FMP according to the timing of IS initiation. Diverse administration patterns were observed in the IS(+) group: immediate IS initiation, comprising patients administered IS immediately upon admission due to the severity of their symptoms; post-EMB IS initiation, comprising patients administered IS only after EMB results indicated a need for immunosuppressive intervention; delayed IS introduction, comprising patients who did not initially receive IS, but a deterioration in their condition necessitated its introduction at a later stage; and discontinued IS treatment, comprising patients who started IS but had to cease its use owing to complications, such as infectious diseases. Given the multicenter nature of our study, variations in treatment strategies across centers were also observed. Although some centers were proactive in initiating IS, others demonstrated a more conservative approach. The intricate patterns in the IS(+) group underscore the multifaceted strategies and outcomes associated with IS administration in patients with FMP. Notably, while the crude analysis revealed pronounced differences in both 90-day and long-term outcomes between the IS(+) and IS(-) groups, these disparities diminished, leading to comparable mortality rates between the groups, after adjustments for patient attributes through PSM.

When patients were categorized based on the initiation of IS on the day of admission, the 90-day rate of mortality or HTx was similar between the two groups (29.6% vs. 28.3%). Subsequently, IS was introduced to 88 patients from the second day of hospitalization to the second day post-EMB, either because of EMB findings or for other reasons. The disparity in the 90-day outcomes increased slightly when comparing patients who did and did not receive IS at this stage (36.6% vs. 23.3%). In the subsequent phase, 53 patients began IS treatment three days after EMB. Here, the difference in 90-day outcomes broadened even more when comparing patients who did and did not receive IS at this stage (35.9% vs. 18.8%). These data suggest that a subset of patients did not respond favorably despite the late initiation of IS in an attempt to provide the most comprehensive care during severe deterioration. Such instances may account for the notable differences in mortality observed in the overall unadjusted population.

When patients were categorized based on the number of prognostic factors identified in prior research,^12^ we found that the rate of 90-day mortality or HTx was marginally higher in the IS(+) group than in the IS(-) group among patients with 2, 3, or 4 or more factors; however, this disparity in outcomes was notably significant among patients with one or zero factors. This suggests that IS could potentially compromise the prognosis of low-risk patients with FMP. The data indicate that IS might not only be ineffective in enhancing FMP outcomes, but could also elevate the risk of infections and other related complications, further deteriorating patient health. Given these findings, there may be merit in exercising caution and potentially avoiding the hasty introduction of IS, especially in low-risk patients.

### Study Limitations

Our study had several limitations. First, as this study was retrospective in nature and situated in actual clinical settings, we meticulously analyzed the link between prognosis and the timing of IS introduction and performed prognosis comparisons based on prognostic risk points. We acknowledge we could not fully account for all biases. Second, the registry only provided data on the initiation date of IS, without detailed information of the dosage, treatment duration, or specifics of the regimen. In addition, details regarding the use of other immunosuppressive drugs were not available. While this study is invaluable because it focuses on a large registry exclusively dedicated to patients with pathologically confirmed FMP, the benefits and drawbacks of steroid use for this condition require definitive confirmation through a prospective randomized trial. This recommendation arises because of the inherent limitations in the design of retrospective studies, such as the present study, and even prospective observational studies.

### Conclusions

In this extensive Japanese cohort study examining FMP, IS were more commonly administered in severe cases, particularly in patients with EM or GCM. Although patients who received IS generally exhibited a poorer prognosis than those who did not receive IS, their outcomes were similar when comparing cohorts matched on background factors. Notably, IS not only appears to be ineffective in enhancing outcomes for FMP, but may also have detrimental effects, especially in low-risk patients with FMP.

## Data Availability

Yes, it is available.

## Non-standard abbreviations and acronyms

EM: eosinophilic myocarditis
EMB: endomyocardial biopsy
FMP: fulminant myocarditis presentation
GCM: giant cell myocarditis
HTx: heart transplantation
IS: intravenous steroids
LM: lymphocytic myocarditis
PSM: propensity score matching

## Acknowledgements

The authors thank Yuki Kamada for their help with data preparation. Drs. Kanaoka, Onoue, and Saito conceived the study. Drs. Kanaoka and Saito managed the data, including quality control. Dr. Kawai drafted the manuscript and Drs. Izawa, Yanase, and Yamada contributed substantially to its revision. Drs. Ozaki and Takada contributed to the study setup. Mr. Takahashi was in charge of statistics. All authors agree with the content of the manuscript.

## Supplementary Material

**Supplemental Figure S1.**
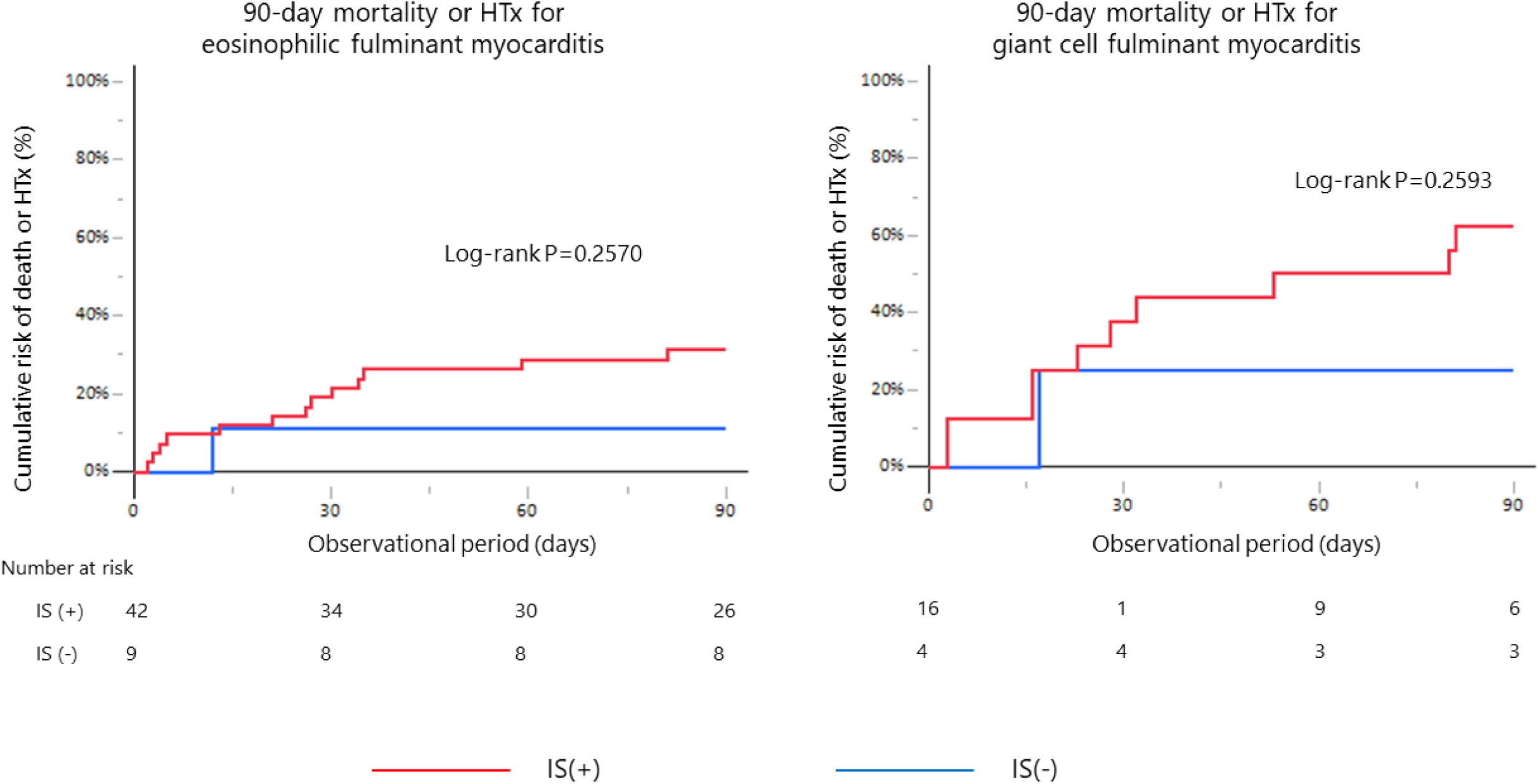
Association of IS use with prognosis in patients with other fulminant myocarditis (a) 90-day mortality or HTx for eosinophilic fulminant myocarditis (b) 90-day mortality or HTx for giant cell fulminant myocarditis IS, intravenous steroid; HTx, heart transplantation

